# Prune intake ameliorates chronic constipation symptoms and causes little discomfort from diarrhea and loose stools: A randomized placebo-controlled trial

**DOI:** 10.1101/2022.01.11.22268876

**Authors:** Taishi Koyama, Naoyoshi Nagata, Kengo Nishiura, Naoki Miura, Takashi Kawai, Hirotaka Yamamoto

## Abstract

**Aim:** Little data is available regarding the effectiveness of natural foods in treating chronic constipation. We aimed to identify whether prune ameliorates chronic constipation and can be used safely for a relatively long time.

**Methods:** In this double-blind, randomized, placebo-controlled trial, 84 subjects with chronic constipation, presenting more than 6 months before and persisting for more than 3 months, were randomized to prune (n=42) or placebo (n=42) intake for 8 weeks. We assessed daily Bristol stool form scale (BSFS) scores and stool frequencies and administered the gastrointestinal symptom rating scale (GSRS) questionnaire, as primary outcomes for constipation improvement.

**Results:** The prune group showed significantly decreased rates of hard stool (BSFS1 or 2) and increased rates of normal stool (BSFS 3 or 4) after 1 week, which were more evident after 7 weeks between the two groups. Prune significantly increased stool frequency immediately after 1 week. Furthermore, GSRS of hard stools, flatulence, and incomplete evacuation significantly improved after 4-8 weeks of prune intake, of which constipation and hard stools were significantly reduced compared to the placebo group. In contrast, prune intake did not cause diarrhea, loose stools, or urgent need for defecation during 8 weeks evaluated by GSRS score. We found no abnormal laboratory tests of liver function, renal function, inflammation, or urinalysis after prune intake.

**Conclusions:** Daily prune intake ameliorates chronic constipation, improving quality of life, and causes few diarrhea-related symptoms or side effects. Our results emphasize a new, useful, and easy strategy for chronic constipation. (UMIN ID:000041384)

## Introduction

Chronic constipation is one of the most frequent digestive disorders, affecting 14% of the world’s population.^1^ Chronic constipation is associated with an increased risk of neurological disease,^2^ cardiovascular disease^3^ and decreased quality of life (QOL), resulting in economic burdens for patients and healthcare providers.^4^ Therefore, there is an urgent need to develop effective and safe treatments for chronic constipation.

Despite the wide variety of laxatives available^5^, patients with chronic constipation may be dissatisfied even under their treatment,^6^ because of side effects including diarrhea, loose stool, and abdominal discomfort,^5^ and insufficient efficacy in reducing constipation and its related QOL.^7^ Coupled with growing public concern regarding the long-term side effects of chronically administered drugs,^8^ constipation therapy has shifted to safer, more natural, non-drug solutions.^9^ Instead of drug treatment, there has been increasing recognition of natural food treatment, which potentially ameliorates chronic constipation.^10^ Natural food treatment is beneficial in its prevalence, safety, low cost, and savings in medical resources.^11^ However, previously there were only 3 randomized controlled trials (RCTs) focusing on the effectiveness of food treatment on chronic constipation.^12–14^

Prunes are dried fruits of a certain type of plum, and are widely consumed in both Western countries and Asian countries.^15^ Because prunes contain a high amount of dietary fiber, sorbitol, and polyphenols,^10^ they are potentially useful in treating constipation.^12^ One RCT showed the effectiveness of prune intake compared to psyllium for chronic constipation,^12^ but the study had some limitations, such as not evaluating symptoms for longer than 4 weeks, a lack of background information concerning dietary, physical, and smoking habits, a possibility of difficulty in continuation due to the relatively high amount of 100g of prunes per day, and insufficient evaluation of lower gastrointestinal symptoms scores other than constipation. Moreover, there are no RCTs conducted in Asian countries focusing on the effects of natural foods for chronic constipation.

To address these issues, we aimed to evaluate the effect of prune consumption on constipation symptoms and lower-gastrointestinal-symptom-related QOL in Japanese individuals with chronic constipation.

## Materials and methods

### Study design, Setting and Participants

The study was conducted under a double-blind, randomized, placebo-controlled design with a 1:1 treatment allocation, and included a minimum 9-week follow-up period (ClinicalTrials.gov number, UMIN ID: 000041384). This study was conducted in accordance with the Declaration of Helsinki. Human rights of the subjects were well protected, and the study was conducted under the supervision of a doctor with the approval of the institutional review board of the Miura Clinic (approval number: R2001) and the Tokyo Medical University (approval number: T2021-0243). Written informed consent was obtained from all subjects prior to their enrollment into the study.

Subjects aged 20-75 years old who presented symptoms of treatment-naive chronic constipation were included in the study. The criterion for chronic constipation was defined with reference to Rome IV as fewer than 3 bowel movements per week, and hard stool was defined by BSFS 1 or 2, for the past 3 months, with symptom onset at least 6 months earlier.^16^ Exclusion criteria included history of diabetes; liver cirrhosis; chronic hepatitis; renal failure; heart failure; cerebral infarction; cerebral hemorrhage; myocardial infarction; angina pectoris; cancer; a history of gastrointestinal surgery (esophagus, stomach, duodenum, small intestine, large intestine, rectum, gallbladder, pancreas); abnormal liver or kidney function within the last 3 months; currently undergoing treatment for a disease; fruit allergies; using acid-secretion inhibitors, antibiotics, or laxatives within the past three months; consuming excessive amounts of alcohol (more than 3 alcoholic drinks/week or more than 50g of alcohol at one time); planning to become pregnant or to breastfeed during the study period; participating in other clinical trials within the last 3 months; judged inappropriate by the principal investigator. To assess eligibility, potential subjects were screened by interview, comprehensive clinical evaluation, and stool investigation using a one week stool diary.

Consent, screening, enrolment and all subsequent study visits took place at the Miura clinic (Osaka, Japan). The study was conducted between July 2020 and January 2021 (from the first informed consent to the last observation).

### Randomization and Masking

Subjects were requested to maintain their normal lifestyles, including diet and physical activity, throughout the study period. After a 1-week baseline period, subjects were randomly assigned to either the prune group or the placebo group. Randomization was by permuted block design, stratified by gender and age. The sequence was generated by a research assistant who was not involved with the current study, using an online random number generator with a 1:1 allocation using fixed block sizes of 4. The allocation sequence was printed and kept in opaque sealed envelopes.

Prune was provided by MIKI Corporation (Osaka, Japan). This is a concentrated type dried plum juice packed in 18g units containing 0.9g of dietary fiber and 2.9g of sorbitol per package as potential active components. The placebo food was 18g units of a packaged concentrate type drink, consisting of equal amounts of sugars (glucose, fructose) as prune, acidulant (malic acid), and a slight amount of flavoring and coloring. Glucose, fructose and malic acid were food-grade ingredients and used because they are originally contained in prune and could make the placebo closer to prune. The placebo food and prune were packaged identically in plain packaging, with similar flavors and colors, in order to make them indistinguishable. Potential active components, dietary fiber, and sorbitol were not contained in the placebo. For an 8-week intake period, subjects were instructed to consume three packages of each test food in a day with some water, and not to consume all three packages at once.

Which group prune was allocated to was blinded to the subjects, research team and statistical analysts involved in the study. Only a few researchers at MIKI Corporation, which created the placebo and prune containers, were left in the dark until the results were available.

### Measurements and follow-up strategy

At baseline, lifestyle factors were assessed, including smoking, alcohol drinking, physical activity, and dietary habits. Stool consistency and stool frequency were recorded in daily stool diaries for a 1-week baseline and an 8-week intake period and analyzed as week by week data. Subjective evaluation of lower-gastrointestinal-symptom-related QOL was measured at baseline 4 weeks, and 8 weeks after starting intake. Compliance was measured during the study period using a daily diary. Subjects were instructed to report any unexpected changes in physical condition to the investigator. Hospital treatment and symptoms deemed serious by the investigator were considered to be adverse events. Safety of the test foods was assessed by laboratory tests conducted at baseline and post-intake period (AST, ALT, ALP, γ-GTP, BUN, Creatinine, Sodium, Potassium, Chloride, CRP and Urinalysis).

### Outcome measures

#### Stool output

Stool consistency and stool frequency were recorded in a daily diary during the 9 weeks of study period. Subjects recorded each bowel movement and its consistency using the Bristol stool form scale (BSFS).^17^ Stool consistency was analyzed as the mean BSFS score, and as the percentage of each score in every week throughout the study period. To evaluate stool frequency, the number of bowel movements in every week was taken from the daily diary throughout the study period.

#### Subjective evaluation of lower-gastrointestinal-symptom-related QOL

Lower-gastrointestinal-symptom-related QOL was assessed using the gastrointestinal symptom rating scale (GSRS) questionnaire.^18^ Seven items regarding lower gastrointestinal symptoms were evaluated: flatulence, constipation, diarrhea, soft stools, hard stools, urgent need for defecation, and incomplete evacuation, using a 7-point Likert scale (1, not present; 2, minor; 3, mild; 4, moderate; 5, moderately severe; 6, severe; and 7, very severe).^19^ The GSRS is a disease-specific 15-item questionnaire developed, based on reviews of gastrointestinal symptoms and clinical experience, to evaluate common symptoms of gastrointestinal disorders. The reliability and validity of the GSRS for functional bowel disease are well-documented.^20^

#### Statistical analysis

The sample size was calculated by considering the effect size estimated from previous studies^21^ on bowel movements. We calculated that 39 subjects per group would be needed to detect a difference in the number of bowel movements per week at a significance level of 0.05 and power of 90%. Assuming a dropout rate of up to 10%, we calculated that 42 subjects per group would have to be enrolled in the study.

Differences in baseline characteristics were assessed using an independent Student’s t-test or a Mann-Whitney U test for continuous variables, or the Chi-square test for categorical variables, respectively. Group differences between the prune and placebo groups during the study period were assessed using the Mann-Whitney U test for continuous variables and the Chi-square test for categorical variables, respectively. Intra-group differences between the baseline and periods from 1 to 8 weeks during the study period were assessed using the paired Wilcoxon signed-rank test for continuous variables, or the McNemar test for categorical variables, respectively.

All data were tested on two-tails, with significance levels of 0.05, and analyzed using IBM SPSS Statistics for Windows, version 20.

## Results

### Baseline characteristics

A total of 354 subjects were screened for eligibility, of whom 84 were randomized (42 in each group) and included in the ITT analysis. During the study period, all subjects were compliant and completed the study without any troubles; therefore we did not make a per-protocol analysis of outcome measures (**Figure 1**). The baseline characteristics of the study population are shown in **Table 1**. There were no significant differences in age, gender, height, weight, BMI, smoking, alcohol, physical activity, and dietary habits between the groups.

**Table 1.**
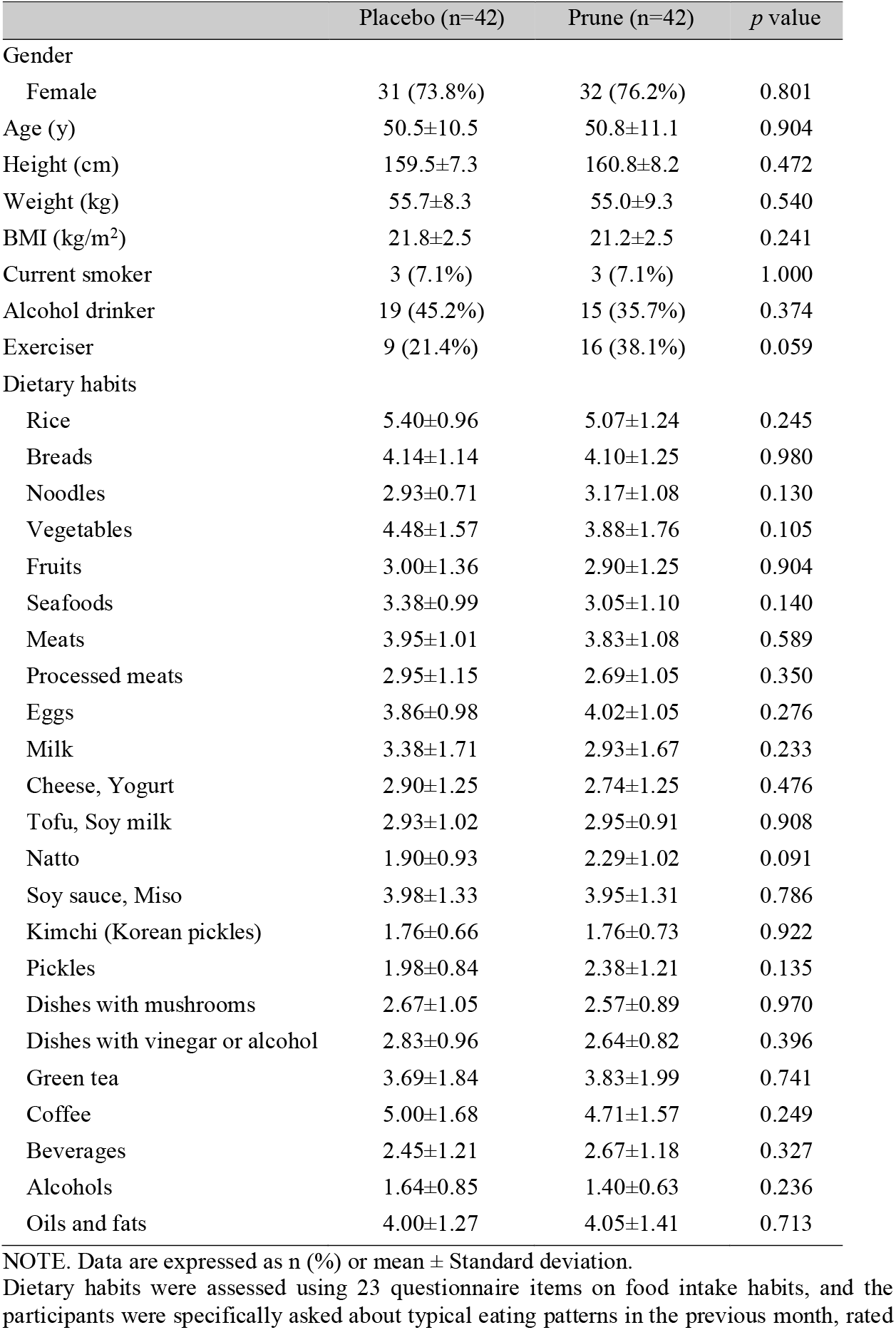
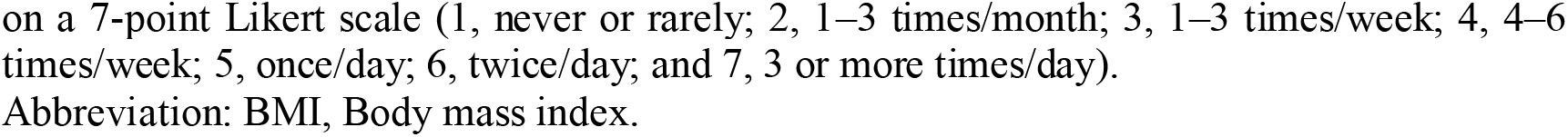
**Baseline characteristics (n=84)**.

|  | Placebo (n=42) | Prune (n=42) | *p* value |
| --- | --- | --- | --- |
| Gender |  |  |  |
| Female | 31 (73.8%) | 32 (76.2%) | 0.801 |
| Age (y) | 50.5±10.5 | 50.8±11.1 | 0.904 |
| Height (cm) | 159.5±7.3 | 160.8±8.2 | 0.472 |
| Weight (kg) | 55.7±8.3 | 55.0±9.3 | 0.540 |
| BMI (kg/m<sup>2</sup>) | 21.8±2.5 | 21.2±2.5 | 0.241 |
| Current smoker | 3 (7.1%) | 3 (7.1%) | 1.000 |
| Alcohol drinker | 19 (45.2%) | 15 (35.7%) | 0.374 |
| Exerciser | 9 (21.4%) | 16 (38.1%) | 0.059 |
| Dietary habits |  |  |  |
| Rice | 5.40±0.96 | 5.07±1.24 | 0.245 |
| Breads | 4.14±1.14 | 4.10±1.25 | 0.980 |
| Noodles | 2.93±0.71 | 3.17±1.08 | 0.130 |
| Vegetables | 4.48±1.57 | 3.88±1.76 | 0.105 |
| Fruits | 3.00±1.36 | 2.90±1.25 | 0.904 |
| Seafoods | 3.38±0.99 | 3.05±1.10 | 0.140 |
| Meats | 3.95±1.01 | 3.83±1.08 | 0.589 |
| Processed meats | 2.95±1.15 | 2.69±1.05 | 0.350 |
| Eggs | 3.86±0.98 | 4.02±1.05 | 0.276 |
| Milk | 3.38±1.71 | 2.93±1.67 | 0.233 |
| Cheese, Yogurt | 2.90±1.25 | 2.74±1.25 | 0.476 |
| Tofu, Soy milk | 2.93±1.02 | 2.95±0.91 | 0.908 |
| Natto | 1.90±0.93 | 2.29±1.02 | 0.091 |
| Soy sauce, Miso | 3.98±1.33 | 3.95±1.31 | 0.786 |
| Kimchi (Korean pickles) | 1.76±0.66 | 1.76±0.73 | 0.922 |
| Pickles | 1.98±0.84 | 2.38±1.21 | 0.135 |
| Dishes with mushrooms | 2.67±1.05 | 2.57±0.89 | 0.970 |
| Dishes with vinegar or alcohol | 2.83±0.96 | 2.64±0.82 | 0.396 |
| Green tea | 3.69±1.84 | 3.83±1.99 | 0.741 |
| Coffee | 5.00±1.68 | 4.71±1.57 | 0.249 |
| Beverages | 2.45±1.21 | 2.67±1.18 | 0.327 |
| Alcohols | 1.64±0.85 | 1.40±0.63 | 0.236 |
| Oils and fats | 4.00±1.27 | 4.05±1.41 | 0.713 |

NOTE. Data are expressed as n (%) or mean ± Standard deviation.
Dietary habits were assessed using 23 questionnaire items on food intake habits, and the participants were specifically asked about typical eating patterns in the previous month, rated
on a 7-point Likert scale (1, never or rarely; 2, 1–3 times/month; 3, 1–3 times/week; 4, 4–6 times/week; 5, once/day; 6, twice/day; and 7, 3 or more times/day). Abbreviation: BMI, Body mass index.

**Figure. 1.**
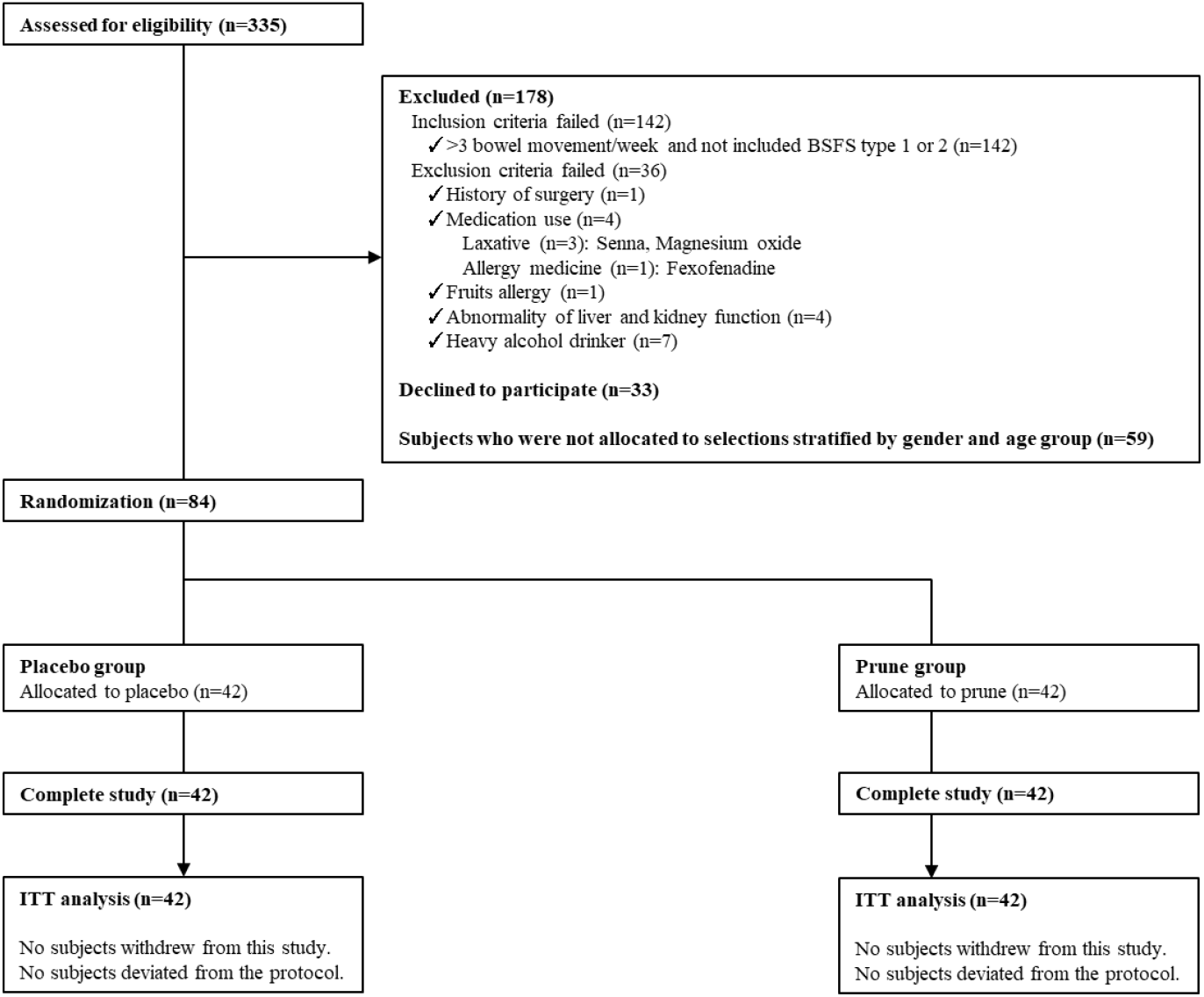
Consort diagram. Abbreviations: BSFS, Bristol stool form scale; ITT, Intention to treat

### Stool consistency

Mean BSFS score during the study period is summarized in **Table 2**. At baseline, mean BSFS scores were not significantly different between the groups, whereas in the prune group, the score increased after 1 week of intake, showing a significant difference from the placebo group after 7 weeks.

**Table 2.**
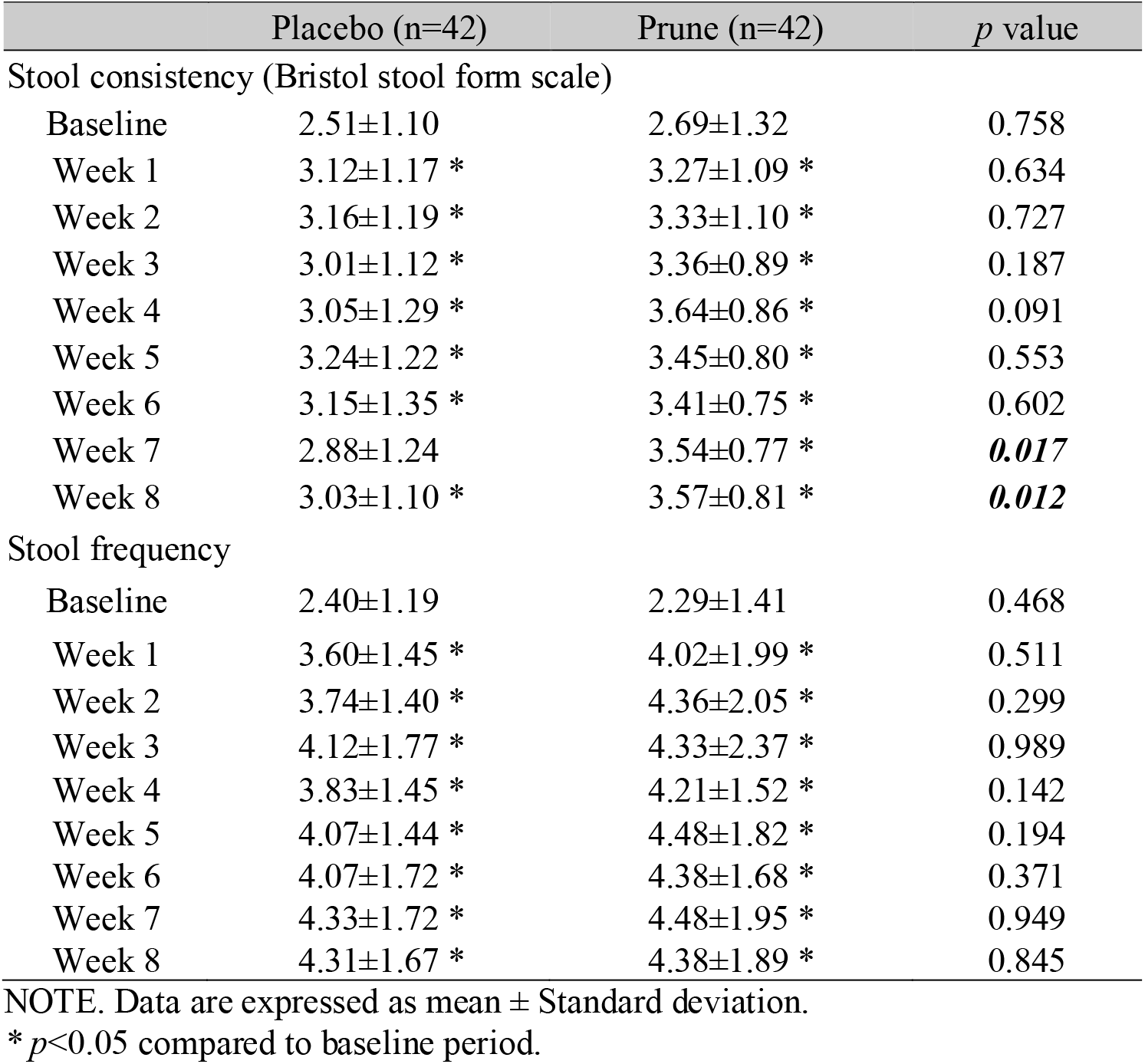
**Mean stool consistency and stool frequency at baseline and intake period (Week 1 to 8)**

Changes in the percentages of each BSFS score are summarized in **Figure 2** and **Supplementary Figure 1**. At baseline, the percentages of all BSFS scores were not significantly different between the groups. In the prune group, the percentage of score 1, indicating the hardest stool, decreased after 3 weeks of intake and showed a significant difference from the placebo group after 3 weeks, except for week 6. A similar tendency was also observed in score 2, indicating the second hardest stool. In contrast, the percentage of score 4, indicating normal stool, increased after 2 weeks of intake, showing a significant difference from the placebo group after 7 weeks. A similar tendency was also observed in score 3, indicating normal but moderately hard stool. The percentages of scores 5, 6, and 7, indicating normal but moderately soft stool to loose stool, were not significantly changed after prune intake.

**Figure 2.**
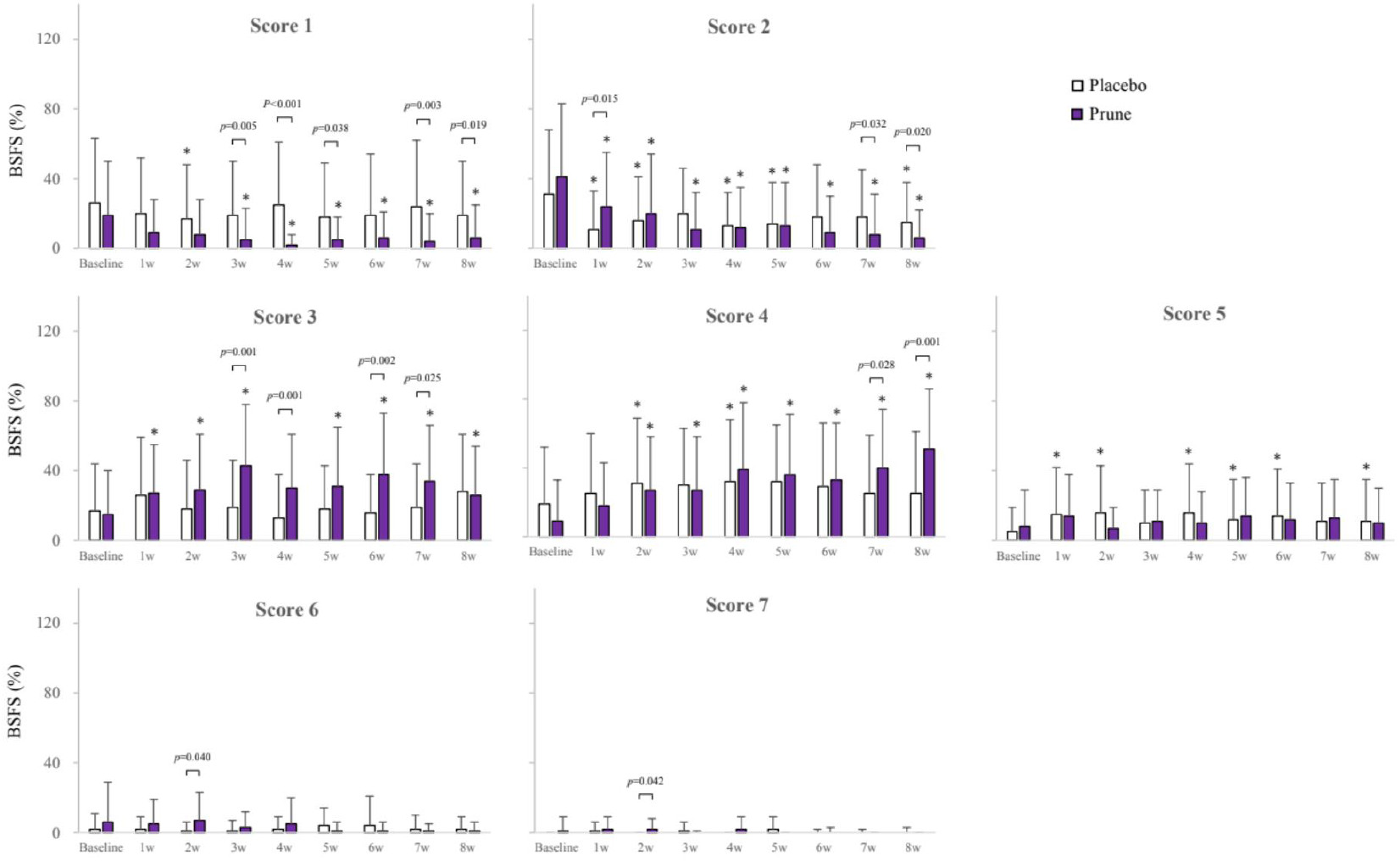
Percentage of each BSFS score at baseline and intake period. Subjects recorded BSFS scores at each bowel movement. Percentage of each BSFS score compared to total bowel movements within 1 week were calculated at baseline and 8 weeks of intake period (W1 to W8). Data are expressed as mean value. Error bars represent standard deviation. n=42 in each group. *p* values in the graph field represent significant differences between the groups. * *p*<0.05 compared to baseline period.

### Stool frequency

At baseline, stool frequency was not different between the groups, whereas in the prune group, stool frequency increased after 1 week of intake (**Table 2**). Stool frequencies represented a higher tendency in the prune group than placebo group at each week, but a significant difference between the groups was not observed at each week.

### Lower gastrointestinal symptoms score with QOL and laboratory abnormalities

At baseline, GSRS scores were not different between the groups in any of the 7 items (**Table 3**). In the prune group, the scores improved in terms of constipation, hard stools, and incomplete evacuation at weeks 4 and 8, and in terms of flatulence at week 8 (**Table 3**). Significant differences from the placebo group were observed in terms of constipation and hard stools at week 8. Meanwhile, the scores in terms of diarrhea, loose stools, and urgent need for defecation were not changed after prune intake and showed no significant difference from the placebo group.

**Table 3.** **Subjective evaluation of gastrointestinal-symptom-related QOL by GSRS questionnaire at baseline and changes from baseline at intake period (Week 4 and 8)**

|  |  | Placebo (n=42) | Prune (n=42) | *p* value |
| --- | --- | --- | --- | --- |
| Flatulence | Baseline | 2.93±1.11 | 3.02±1.24 | 0.801 |
|  | Δ4w | -0.36±1.21 * | -0.48±1.35 | 0.480 |
|  | Δ8w | -0.38±1.13 | -0.43±1.36 * | 0.982 |
| Constipation | Baseline | 4.50±0.99 | 4.74±1.01 | 0.490 |
|  | Δ4w | -1.24±1.41 * | -1.62±1.36 * | 0.163 |
|  | Δ8w | -1.43±1.56 * | -2.24±1.41 * | **0.024** |
| Diarrhea | Baseline | 1.55±0.80 | 1.57±0.83 | 0.826 |
|  | Δ4w | 0.14±1.20 | 0.07±1.09 | 0.914 |
|  | Δ8w | 0.05±0.94 | -0.05±1.08 | 0.741 |
| Loose stools | Baseline | 1.64±0.91 | 1.48±0.59 | 0.714 |
|  | Δ4w | -0.02±1.00 | 0.21±0.90 | 0.352 |
|  | Δ8w | 0.02±0.87 | 0.26±0.91 | 0.412 |
| Hard stools | Baseline | 4.33±1.37 | 4.81±1.13 | 0.172 |
|  | Δ4w | -1.26±1.58 * | -1.83±1.32 * | 0.082 |
|  | Δ8w | -1.33±1.71 * | -2.31±1.30 * | **0.009** |
| Urgent need for defecation | Baseline | 2.38±1.13 | 2.33±1.32 | 0.643 |
|  | Δ4w | 0.05±1.38 | -0.10±1.74 | 0.934 |
|  | Δ8w | -0.26±1.15 | -0.21±1.57 | 0.703 |
| Incomplete evacuation | Baseline | 4.24±1.49 | 4.36±1.36 | 0.938 |
|  | Δ4w | -1.12±1.70 * | -1.48±1.47 * | 0.219 |
|  | Δ8w | -1.45±1.56 * | -1.81±1.42 * | 0.222 |

NOTE. Data are expressed as mean ± Standard deviation.
\* $p < 0.05$ compared to baseline period.
Abbreviations: QOL, Quality of life. GSRS, Gastrointestinal symptoms rating scale.

There were no significant differences of liver function, renal function, inflammation, or urinalysis results before and after prune intake as well as placebo intake (**Table 4**). There were no adverse events during the study period, and no subjects who had visited a hospital for prune or placebo intake during this study.

**Table 4.** **Percentage of subjects with abnormal laboratory tests at baseline and post-intake during the study period**.

|  | Placebo |  |  | Prune |  |  |
| --- | --- | --- | --- | --- | --- | --- |
|  | Baseline<br>(n=42) | Post-intake<br>(n=42) | *p* value | Baseline<br>(n=42) | Post-intake<br>(n=41)<sup>†</sup> | *p* value |
| AST (U/L) | 0 (0.0%) | 1 (2.4%) | 1.000 | 0 (0.0%) | 1 (2.4%) | 1.000 |
| ALT (U/L) | 0 (0.0%) | 2 (4.8%) | 0.500 | 0 (0.0%) | 1 (2.4%) | 1.000 |
| ALP (U/L) | 1 (2.4%) | 1 (2.4%) | 1.000 | 1 (2.4%) | 0 (0.0%) | 1.000 |
| γ-GTP (U/L) | 2 (4.8%) | 4 (9.5%) | 0.500 | 1 (2.4%) | 1 (2.4%) | 1.000 |
| Creatinine (mg/dL) | 1 (2.4%) | 2 (4.8%) | 1.000 | 0 (0.0%) | 0 (0.0%) | - |
| BUN (mg/dL) | 1 (2.4%) | 0 (0.0%) | 1.000 | 3 (7.1%) | 4 (9.8%) | 0.625 |
| Sodium (mEq/L) | 0 (0.0%) | 0 (0.0%) | - | 0 (0.0%) | 0 (0.0%) | - |
| Potassium (mEq/L) | 0 (0.0%) | 0 (0.0%) | - | 1 (2.4%) | 1 (2.4%) | 1.000 |
| Chloride (mEq/L) | 0 (0.0%) | 0 (0.0%) | - | 0 (0.0%) | 0 (0.0%) | - |
| CRP (mg/dL) | 0 (0.0%) | 3 (7.1%) | 0.250 | 0 (0.0%) | 1 (2.4%) | 1.000 |
| Protein in Urine | 2 (4.8%) | 2 (4.8%) | 1.000 | 0 (0.0%) | 0 (0.0%) | - |
| Sugar in Urine | 0 (0.0%) | 0 (0.0%) | - | 0 (0.0%) | 0 (0.0%) | - |
| Urobilinogen in Urine | 0 (0.0%) | 0 (0.0%) | - | 1 (2.4%) | 0 (0.0%) | 1.000 |
| Bilirubin in Urine | 0 (0.0%) | 0 (0.0%) | - | 0 (0.0%) | 0 (0.0%) | - |
| Urine pH | 0 (0.0%) | 0 (0.0%) | - | 0 (0.0%) | 0 (0.0%) | - |
| Occult Blood in Urine | 3 (7.1%) | 1 (2.4%) | 0.688 | 6 (14.3%) | 6 (14.6%) | 1.000 |
| Urine Specific Gravity | 0 (0.0%) | 0 (0.0%) | - | 0 (0.0%) | 0 (0.0%) | - |
| Ketone in Urine | 0 (0.0%) | 0 (0.0%) | - | 1 (2.4%) | 1 (2.4%) | 1.000 |

NOTE. Data are expressed as n (%) who showed abnormal value.
Abnormal range defined as >1.2x upper limit of normal range and <1.2x lower limit of normal range.
<sup>†</sup> One subject withdrew from comprehensive clinical evaluation.
Abbreviations: AST, Aspartate aminotransferase. ALT, Alanine aminotransferase. ALP, Alkaline Phosphatase. γ-GTP, γ-Glutamyl transpeptidase.
BUN, Blood urea nitrogen. CRP, C-reactive protein.

## Discussion

This randomized double-blind placebo control study first focused on the effects of prune on chronic constipation. First, in the prune group, the mean BSFS score significantly increased after 1 week of intake, showing a significant difference from the placebo group after 7 weeks (**Table 2**). Intriguingly, the prune group showed significantly decreased rates of hard stool (BSFS1 or 2) after 1 week of intake, and increased rates of normal stool (BSFS 3 or 4) after 1 week (**Figure 2**), becoming more evident after 7 weeks (**Figure 2**). Moreover, prune significantly increased stool frequency immediately after 1 week (**Table 2**). Second, QOL scores for hard stools, flatulence, and incomplete evacuation significantly improved after 4 and 8 weeks of prune intake, of which constipation and hard stools were significantly pronounced compared to the placebo group (**Table 3**). Third, prune intake did not cause diarrhea, loose stools, or urgent need for defecation during the 8 weeks evaluated by GSRS score (**Table 3**). Furthermore, there were no significant differences in abnormal laboratory tests of liver function, renal function, inflammation, or urinalysis between times before and after prune intake (**Table 4**). These findings highlight an ameliorative effect and the safety of prune intake to address chronic constipation symptoms.

We conducted a detailed, objective, longitudinal evaluation of bowel symptoms with BSFS and GSRS. Interestingly, the improvement of constipation symptoms by BSFS was consistent with the improvement of subjects’ own constipation symptom QOL by GSRS, which strongly suggests that prunes not only change the actual shape of stools, but also provide a high level of satisfaction with the improved effect. In contrast, prunes are known to cause symptoms such as diarrhea^22^ and flatulence,^23^ and the key to adhering to prune intake is the low frequency of these symptoms. Considering that prunes do not cause abnormal findings in laboratory tests suggests that prunes are a continuous and easy-to-use treatment for constipation. Ohkubo et al. reported that the persistence of the percentage of BSFS score 4 correlated with the QOL as assessed by the PAC score.^24^ In the present study, the percentage of score 4 increased significantly after 7 weeks of prune consumption compared to placebo. These facts suggest that the effect of prune intake on constipation should not be evaluated in a short period of time, but should be continued for at least nearly 2 months. Previous studies failed to show the increased percentage of score4 because of the 4-week evaluation^12^, but we were able to obtain this because of the 8-week long-term evaluation.

The stool frequency was significantly increased after 1 week to 8 weeks of prune intake compared to baseline period, but, as the placebo group also showed increased stool frequency, there was no significant difference between the groups (**Figure 2**). A previous study by Attaluri et al.^12^ observed a significant increase in stool frequency after 3 weeks of prune intake, and a significant difference from its psyllium group. One possible reason for the observed differences could be the placebo effect. A number of studies using chronic constipation patients have reported a significant increase in stool frequency in the placebo group.^25,26^ The use of a clear placebo in the present study may have led to an increase in the stool frequency in the placebo group, masking the effect of the prune on stool frequency. Another possible reason might be due to lower intake of prune in our study.

In the present study, we provided 54g of prunes daily, approximately half the amount used in the previous RCT showing that the intake of 100g of prunes for 4 weeks significantly improved constipation.^12^ It is estimated that 100g of daily prune is equivalent to 12 prunes,^12^ suggesting that large daily intake of natural foods poses a risk of diminishing continuous adherence. Moreover, it has been reported that prunes have abundant amounts of glucose (23g/100g) and fructose (13g/100g).^10^ Excessive daily intake of these free sugars has recently been reported to be a risk factor for non-communicable diseases such as cancer, diabetes, and cardiovascular disease.^27^ We are concerned about the potential difficulty in consuming this amount on a consistent daily basis and the risk of increased blood sugar associated with the amount.

The strength of our study is that we have investigated detailed background factors potentially associated with constipation, such as dietary habits, smoking, physical activity, and alcohol consumption,^28^ which were lacking in previous studies.^12^ 8 weeks of evaluation of BSFS, frequencies, GSRS, and laboratory data associated with food intake enable us to conclude the efficacy and safety of prunes for a long period. In contrast, a limitation of our study is that we did not evaluate the palatability of the test foods. Moreover, subjects in our study were not using laxatives. It remains unknown whether laxatives with prune are effective and safe for chronic constipation.

In conclusion, this study demonstrated that prune intake ameliorates chronic constipation symptoms and causes little discomfort from diarrhea and loose stools in Japanese individuals. The results indicate that easy to take prunes, which are available worldwide, are effective for treating chronic constipation. Assessment at more acceptable doses and in various ethnic groups would contribute to expansion of our understanding of prune as a natural food treatment for chronic constipation.

## Supporting information

CONSORT check list for randomized trial

## Data Availability

All data produced in the present study are available upon reasonable request to the authors.

## Acknowledgements

We thank Masashi Nakagawa, Humihiro Shimokawa, and Hideki Watanabe for their support in planning this study. The food products used in the study were provided by MIKI Corporation. This study was funded in full by MIKI Corporation. Thane Doss contributed to editing of the manuscript.

