## Supplementary figure 1 for "Prune intake ameliorates chronic constipation symptoms and causes little discomfort from diarrhea and loose stools: A randomized placebo-controlled trial"

### Slide 1
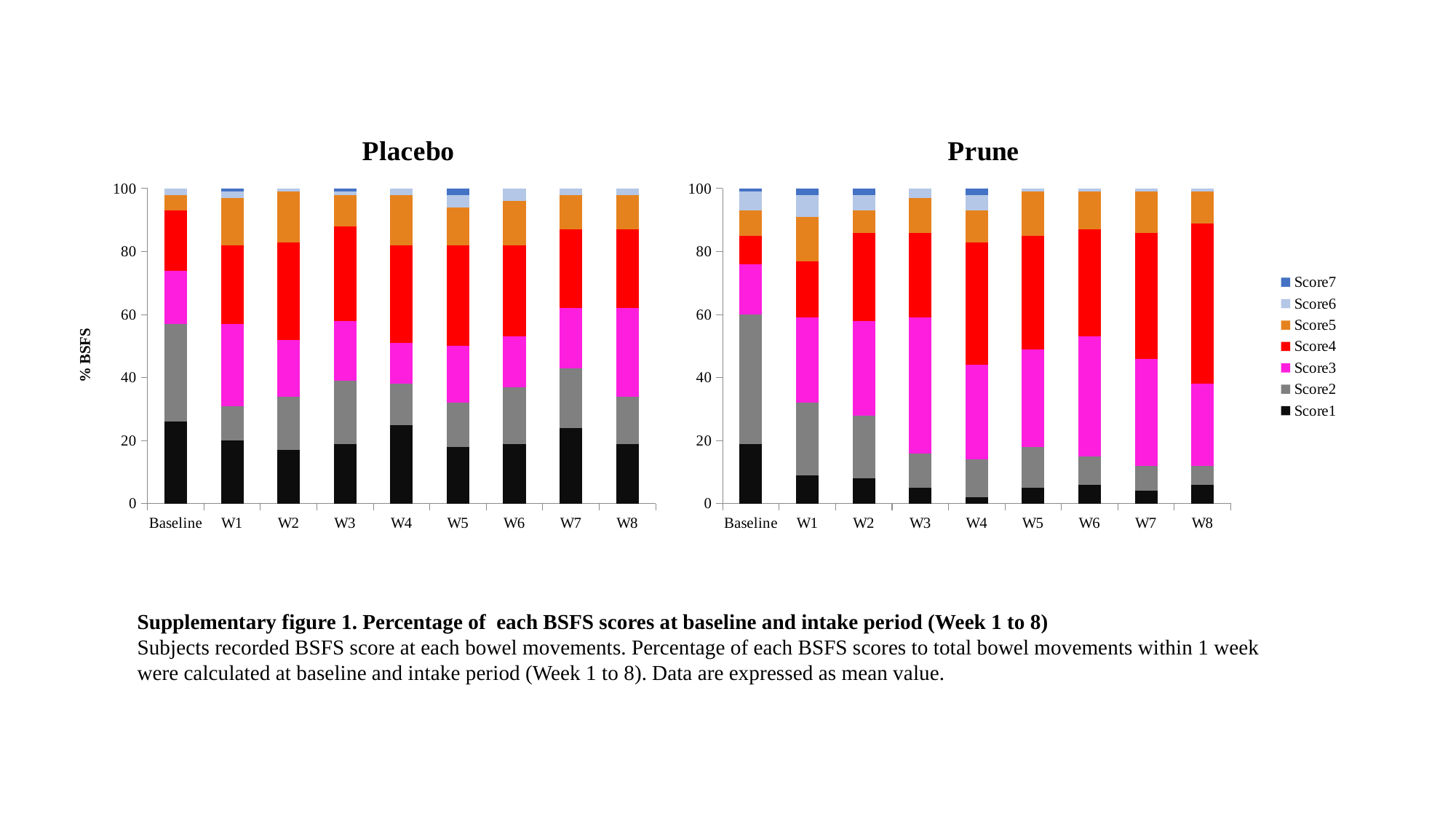

#### Chart: Placebo
| Category | Type1 | Type2 | Type3 | Type4 | Type5 | Type6 | Type7 |
|---|---|---|---|---|---|---|---|
| Baseline | 26.0 | 31.0 | 17.0 | 19.0 | 5.0 | 2.0 | 0.0 |
| W1 | 20.0 | 11.0 | 26.0 | 25.0 | 15.0 | 2.0 | 1.0 |
| W2 | 17.0 | 17.0 | 18.0 | 31.0 | 16.0 | 1.0 | 0.0 |
| W3 | 19.0 | 20.0 | 19.0 | 30.0 | 10.0 | 1.0 | 1.0 |
| W4 | 25.0 | 13.0 | 13.0 | 31.0 | 16.0 | 2.0 | 0.0 |
| W5 | 18.0 | 14.0 | 18.0 | 32.0 | 12.0 | 4.0 | 2.0 |
| W6 | 19.0 | 18.0 | 16.0 | 29.0 | 14.0 | 4.0 | 0.0 |
| W7 | 24.0 | 19.0 | 19.0 | 25.0 | 11.0 | 2.0 | 0.0 |
| W8 | 19.0 | 15.0 | 28.0 | 25.0 | 11.0 | 2.0 | 0.0 |
#### Chart: Prune
| Category | Score1 | Score2 | Score3 | Score4 | Score5 | Score6 | Score7 |
|---|---|---|---|---|---|---|---|
| Baseline | 19.0 | 41.0 | 16.0 | 9.0 | 8.0 | 6.0 | 1.0 |
| W1 | 9.0 | 23.0 | 27.0 | 18.0 | 14.0 | 7.0 | 2.0 |
| W2 | 8.0 | 20.0 | 30.0 | 28.0 | 7.0 | 5.0 | 2.0 |
| W3 | 5.0 | 11.0 | 43.0 | 27.0 | 11.0 | 3.0 | 0.0 |
| W4 | 2.0 | 12.0 | 30.0 | 39.0 | 10.0 | 5.0 | 2.0 |
| W5 | 5.0 | 13.0 | 31.0 | 36.0 | 14.0 | 1.0 | 0.0 |
| W6 | 6.0 | 9.0 | 38.0 | 34.0 | 12.0 | 1.0 | 0.0 |
| W7 | 4.0 | 8.0 | 34.0 | 40.0 | 13.0 | 1.0 | 0.0 |
| W8 | 6.0 | 6.0 | 26.0 | 51.0 | 10.0 | 1.0 | 0.0 |Supplementary figure 1. Percentage of each BSFS scores at baseline and intake period (Week 1 to 8)
Subjects recorded BSFS score at each bowel movements. Percentage of each BSFS scores to total bowel movements within 1 week were calculated at baseline and intake period (Week 1 to 8). Data are expressed as mean value.
